# The Changing Landscape: Geospatial, Epidemiologic Characteristics and Infectious Diseases of Inmates Living in a Region with a High Number of Prisons, Brazil

**DOI:** 10.1101/2020.01.10.20016998

**Authors:** Charlene Troiani do Nascimento, Danilo Zangirolami Pena, Rogério Giuffrida, Fernanda Nobre Bandeira Monteiro, Francisco Assis da Silva, Edilson Ferreira Flores, Luiz Euribel Prestes Carneiro

## Abstract

**Objective:** We determined the geospatial and epidemiologic characteristics and prevalence of HIV, tuberculosis, viral hepatitis, syphilis, and co-infections in inmates in 28 prisons.

**Design:** This is a regional, observational, retrospective, and descriptive study conducted from November 2017 to October 2018.

**Setting:** Prisons are located in the western and northwestern regions of São Paulo state, Brazil.

**Methods:** Data were obtained through a standard questionnaire from inmates with a diagnosis of infectious diseases and co-infections (HIV, hepatitis B virus, hepatitis C virus, syphilis, and tuberculosis), treatment, and time of incarceration. Locations of prisons and highways were obtained from shape file databases. Data on inhabitants, population growth, and Human Development Index were obtained from public agencies. Maps were constructed using geographic information system.

**Results:** A total of 37,497 individuals were enrolled in the study and 741 (1.97%) were diagnosed. HIV was the most prevalent disease (0.68%), followed by tuberculosis (0.66%), syphilis (0.2%), HCV (0.2%), and HBV (0.04%). HIV-syphilis was the most prevalent co-infection (odds ratio, 63.7; 95% confidence interval: 41.4, 96.7). There was a statistical significance (*P*<0.001) for those with HIV acquiring co-infections. In 21 units, there was no relationship between the number of prisoners and the prevalence of infectious diseases.Mean age was 35.82 years (SD, 10.41 years) and 57.9% had been in prison previously. Strategically, most prisons were constructed beside radial highways. A higher than expected population growth was observed in 17 municipalities in which prisons were constructed (47.2%).

**Conclusions:** This is one of the biggest studies in Brazil, and the prevalence of infectious diseases among inmates was lower than countrywide. Therefore, improvements in health care are necessary, mainly in screening for infectious diseases. Construction of large prisons beside the radial highways changed the landscape and prevented migration of people from small to large cities.

**Strengths and limitations of this study:**

- The western and northwestern regions of São Paulo state have the highest number of prisons in Brazil and the risk of infectious diseases in prisoners is higher than that in the general population.
- Few studies have addressed this issue countrywide and for this reason, we determined the prevalence of HIV, HBV, HCV, syphilis, tuberculosis and co-infections as well as epidemiologic characteristics in 37,497 inmates of 28 prisons.
- The data were obtained through a questionnaire sent to each prison health care unit. Due to lack of commitment to complete the questionaire and as a retrospective study, some important data cannot be measured and therefore are limitations of this study.
- We used geospatial analytic techniques to understand the geographic strategy used for the location and construction of the prisons.
- Most prisons were constructed in small-sized cities. We used the number of inhabitants, population growth and human developing index to analyze their impact on preventing migration and development in the municipalities in which prisons were constructed.

## INTRODUCTION

The risk of an infectious disease in a prison is higher than in the general population and can be triggered by different factors, including structural and logistic characteristics of prisons and by behavioral habits acquired before or during imprisonment. [1] However, the prison should provide inmates with access to the public health system, including vaccine programs, diagnosis, and treatment especially of infectious diseases. [2, 3] Furthermore, prisons can help to decrease dissemination, especially to inmates’ partners. [1]

The number of inmates is increasing worldwide, including in Brazil, with a 707% increase in 2016 since the 1990s. Brazil has the third largest prison population in the world, with 726,712 individuals in 1,422 facilities and an occupancy rate of 197.4%. São Paulo state had the biggest prison population with 240,061 inmates and an occupancy rate of 183%, representing 33.0% of inmates countrywide. [4] The western and northwestern regions have a high number of prisons with 39 units in 2018, housing an estimated 57,700 individuals; 80% of the units are overcrowded. [5] All prisons are coordinated by Coordination of Prison Units of Western Region (CROESTE). This situation, together with other risk factors, makes inmates vulnerable to several infectious diseases such as HIV, viral hepatitis, tuberculosis (TB), and syphilis.

The western and northwestern regions of São Paulo state are characterized by small municipalities, most with a population <20,000 inhabitants. In contrast to dynamic economic situation in the rest of the state, sustained growth is hampered by ongoing conflicts between landowners and the rural workers’ movement (MST). [6] Since 1997, to cover the shortage of prisons in the country and the collapse of the penitentiary system, several prisons have been constructed in the region with two main objectives: (i) decentralization of prisons from the capital São Paulo to country areas; (ii) to increase the regional economy and development.

Breaking the monotony of cattle pastures and sugar cane plantations, a changing landscape is observed by people who travel on the highways of the region. Travelers are now confronted with large prisons, mainly maximum security penitentiaries. However, there are many questions at state and regional levels that have not yet been sufficiently addressed: (i) although there is a standard protocol for screening infectious diseases, what is the prevalence in prisons of CROESTE; (ii) have the increasing number of prisons prevented migration of the local population to larger cities such as São Paulo and increased the Human Development Index (HDI) of the local population and the number of jobs on offer; (iii) was a geographic strategy criteria used to choose the location of prisons?

There are few studies highlighting infectious diseases and geospatial characteristics of inmates in prisons in São Paulo state and none for the western and northwestern regions. [7] Our objective was to determine the geospatial, epidemiologic characteristics and prevalence of infectious diseases in inmates in a region with a high number of prisons in São Paulo state.

## METHODS

### Study design and regional characteristics

São Paulo state, the richest and most populous state of Brazil, is located in the southeastern region and includes 645 municipalities (Figure 1A). In June 2019, the state has 173 prisons; in 2018, there were 39 units in the western and northwestern region under the supervision of CROESTE (Figure 1B). The region covers a large area bordering Mato Grosso do Sul, Minas Gerais, and Paraná states. The western region, with an estimated population of 900,000, is considered the second poorest region in the state (Figure 1B). In 1997, to cover the deficit in the country’s prison system and the collapse of the penitentiary system, the Federal Government launched a countrywide program for the construction of new prisons. At that time, São Paulo had the largest prison population in the country, with a great shortage of penal establishments. [8] Since 1997, an increasing number of units have been constructed mainly along the highways linking the bordering states of Mato Grosso do Sul, Minas Gerais, and Paraná (Figure 1C).

**Figure 1.**
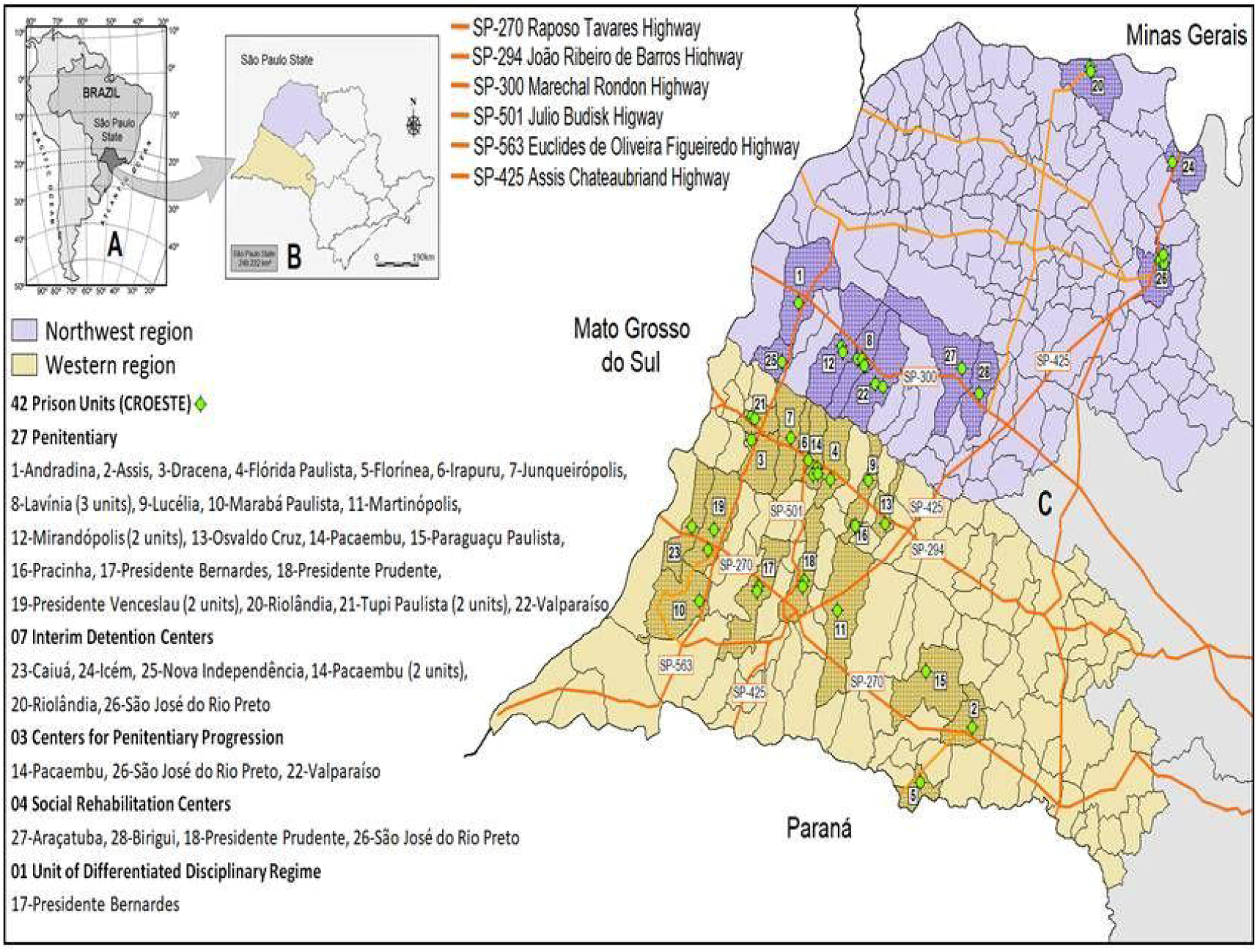
Geospatial location of São Paulo state, regions with prisons, and road connections. This map shows the regions and municipalities where the prisons were constructed, the extensive radial and transverse highways network linking the municipalities to Presidente Prudente (number 18) and São Paulo. Current shape file databases and base maps were downloaded from the IBGE website.

This is an observational, retrospective, and descriptive study conducted from November 2017 to October 2018 on inmates in prisons under supervision of CROESTE (Figure 1B). Of the 39 prisons, with an estimated population of 57,700 individuals, 28 (71.8%) representing 37,497 individuals took part in the study.

### Ethical approval

All procedures performed in studies involving participating human subjects were in accordance with the ethical standards of the institution and/or national research committee and with the 1964 Helsinki Conference declaration and its subsequent amendments or comparable ethical standards. This study was approved by the Ethics Committee of Oeste Paulista University (protocol number CAAE 73092117.1.0000.5515 CPDI 4088) and by the Ethics Committee of Public Security Secretary of São Paulo state (protocol number 2.639.635). As a retrospective and observational study that dispenses the collection of direct information with the research subject, the waiver of the Informed Consent Form was approved by the Ethics Committee of Public Security Secretary of São Paulo state.

### Questionnaire

In each prison, the health care team completed a standard questionnaire, based on data obtained from record files. The questionnaire included epidemiologic information: age, gender, schooling, sexual behavior, marital status, number of sons, diagnosis of infectious diseases and co-infections (HIV, hepatitis B virus [HBV], hepatitis C virus [HCV], syphilis, and TB, treatment, and time of incarceration. Male and female inmates’ ≥18 years of age were included. Laboratory diagnosis followed the protocols of the Brazilian Ministry of Health. [9-12]

### Prevalence of infectious diseases in prisons in western São Paulo state

Although there is a standard state protocol for screening infectious diseases in prisons, we wanted to know if there was a relationship between the number of inmates and the prevalence of infectious diseases in CROESTE prisons. The proportions of infected individuals in each prison were estimated and the 95% confidence intervals (CI) were obtained by Wilson’s method. [13] A Forest plot was constructed to show the increasing order of prevalence (Figure 2).

**Figure 2.**
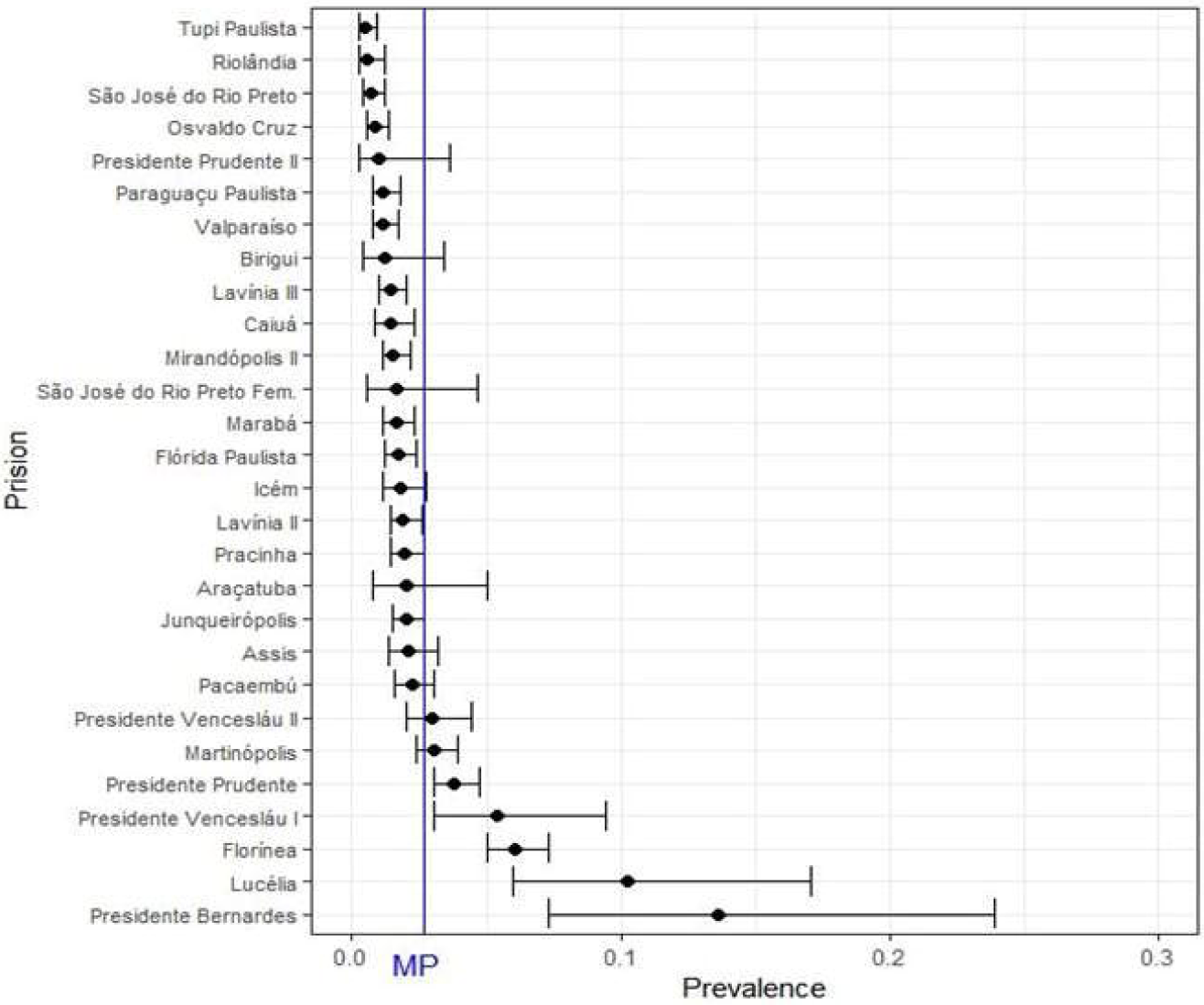
Forest plot for prevalence estimates of infectious diseases in prison inmates in the western and northwestern region of São Paulo state. MP, mean prevalence.

The association between HIV positivity and co-infections was evaluated by estimated odds ratios (ORs) and 95% CI. [14]

### Highways network and geospatial location of prisons

To understand the geographic strategy used for construction of prisons in the CROESTE domain, we plotted their location along the highways network of western and northwestern São Paulo state, [15] categorized as radial roads (main roads connecting municipalities to the state capital) and transverse highways (smaller roads, including local highways connecting municipalities). Current shape file databases and base maps were downloaded from the IBGE (Instituto Brasileiro de Geografia e Estatística) website.

### Inmates by prison and infected individuals

The map showing the number of inmates in each prison was created in ArcGIS software 10.2.2 (ESRI, Redlands, CA), using the Symbology-Show Quantiles tool from Layer Properties. Five classes were generated manually at equal intervals. The variable represented was the total number of prisoners. The Symbology-Show Charts Bar tool was used to present cases of infectious diseases in columns (white bars). Colors represent the number of inmates in each municipality.

### Number of inhabitants, population growth, and Human Development Index of municipalities with prisons

To analyze if the construction of prisons in the region prevented migration of the local population and led to an increase in the HDI of municipalities, we compared the population in the year the prisons were constructed with the expected population in 2018. The data were obtained from the IBGE and State Database System (SEADE). The estimated population growth was obtained by multiplying the population in the year that the prison was constructed by 1.3 (annual growth from the 2000-2010 census) [16] and the number of years to 2018. The HDI was used to analyze the socioeconomic situation of the municipalities in the CROESTE domain.

### Statistical analysis

Results are shown as mean ± standard error of the mean (SEM) (if the variable is normally distributed) and median (interquartile range) for continuous variables. The association between qualitative variables was assessed using the odds ratio (OR) with 95% confidence intervals (95% CI). Dichotomous and nominal variables were expressed as frequencies and percentages. Statistical analysis was performed using the GraphPad Software (San Diego, CA, USA) and Sigma-Stat program (Systat Software Inc., Richmond, CA, USA).

## RESULTS

### Prevalence of infectious diseases and co-infections

Of the 37,497 inmates included in the study, 741 (1.97%) were diagnosed with at least one of the diseases and co-infections investigated. The prevalence of HIV was 0.68% (n=256), TB 0.66% (n=247), syphilis 0.2% (n=75), HCV 0.17% (n=65), HBV 0.04% (n=16), and co-infection 0.22% (n=82). Three (0. 4%) of those diagnosed with an infection were female (Table 1). The number of patients treated was significantly higher compared with non-treated (*P*<00001, mean, 0.01157; 95% CI: 0.008241, 0.01624, with Woolf approximation).

**Table 1.**
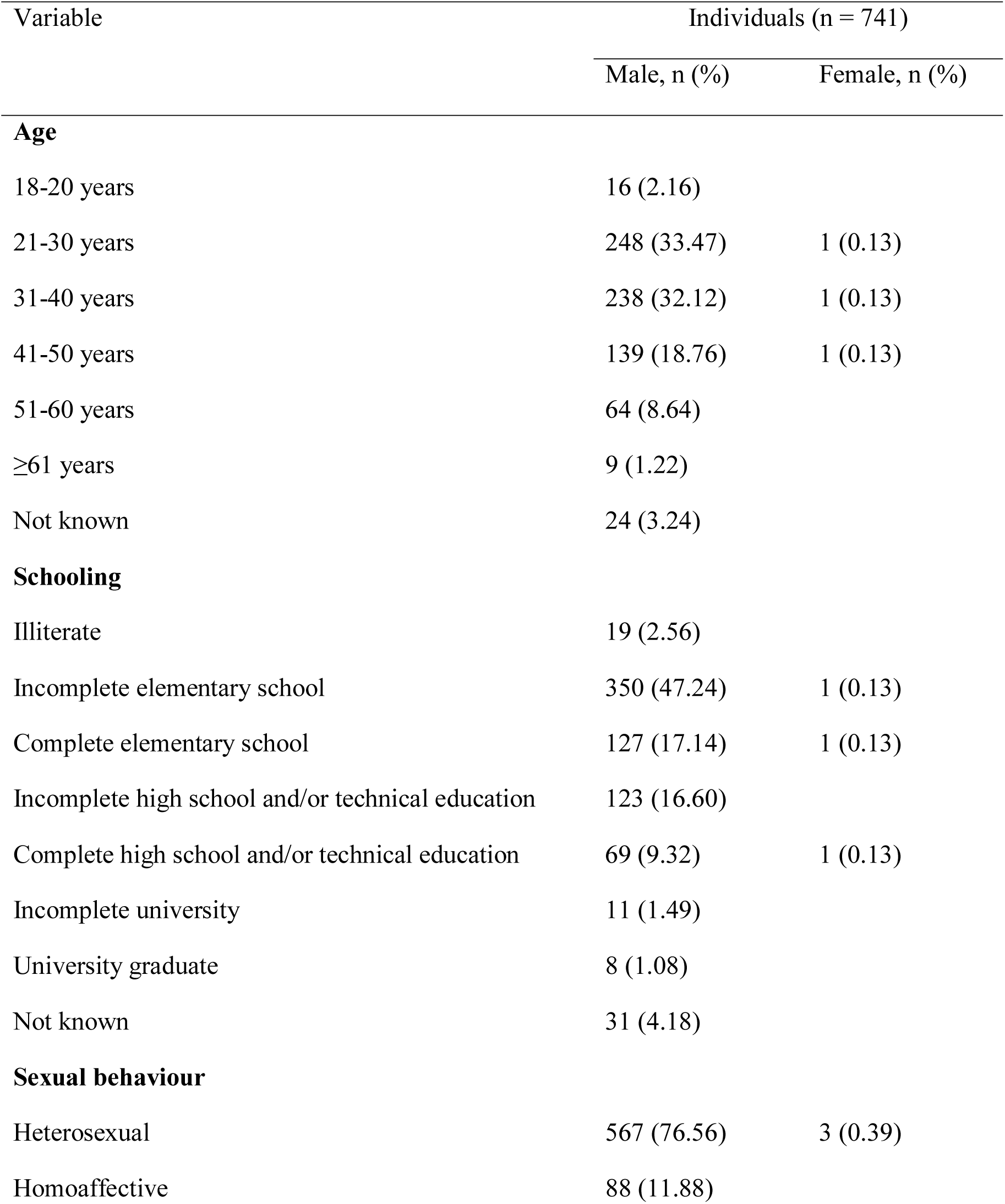
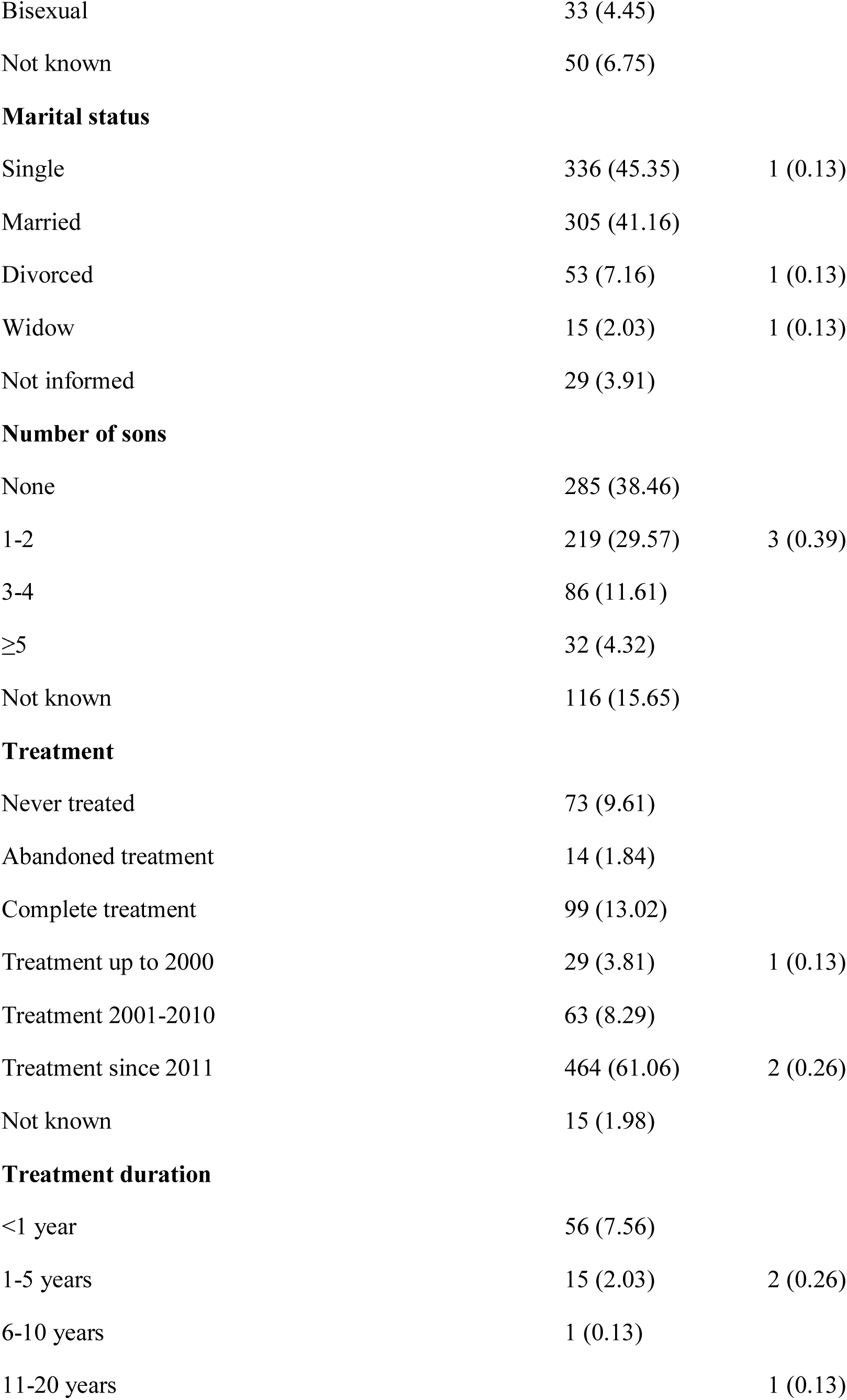
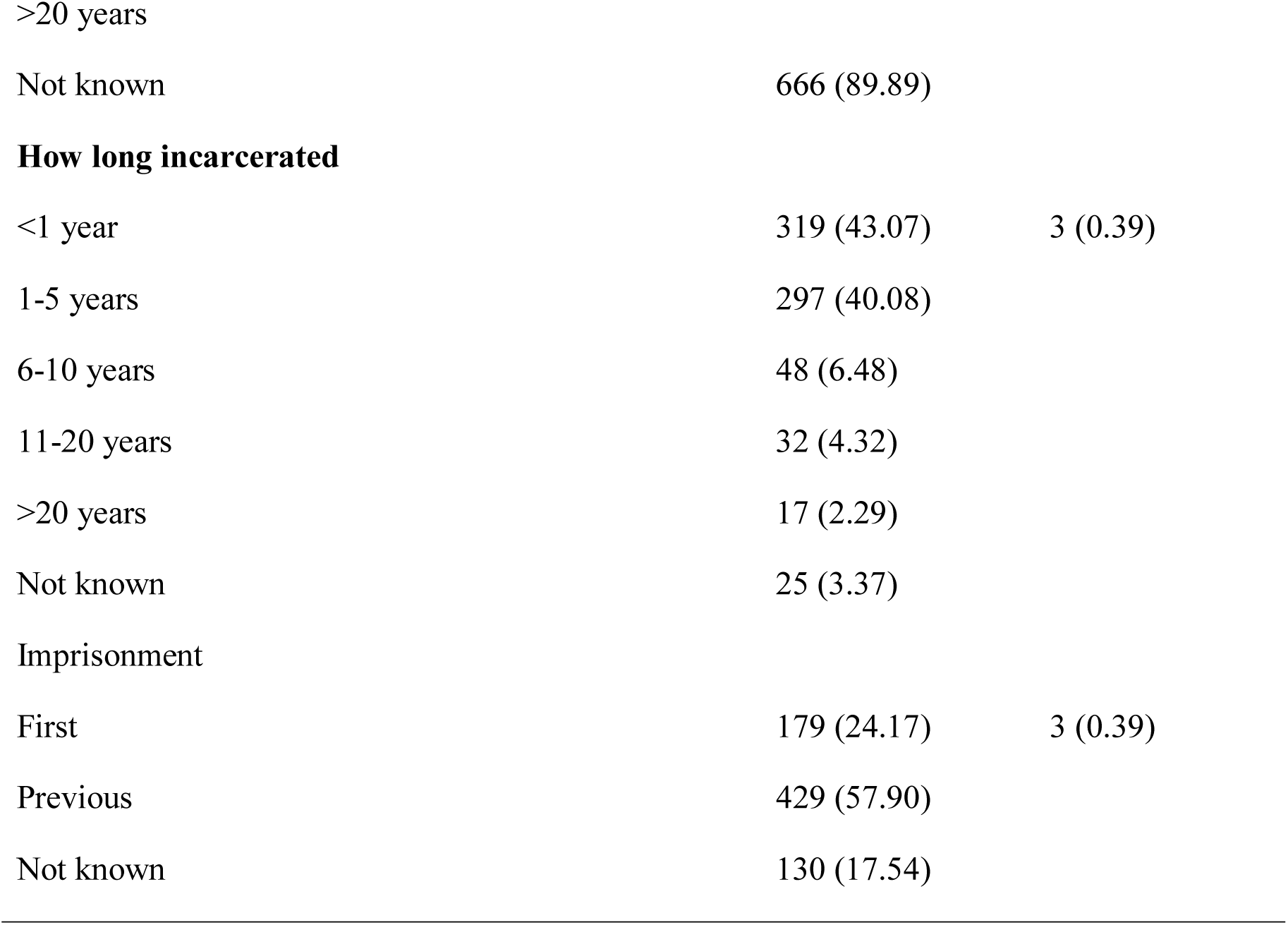
Clinical, Epidemiologic Characteristics and related to the Treatment of Infectious Diseases and Time of Incarceration of 741 Inmates Diagnosed with Infectious Diseases in Prisons in the Western and Northwestern Regions of São Paulo State, Brazil, 2018

### Epidemiologic characteristics

The mean age of male inmates was 35.82 years (SD 10.41 years), ranging from 19 to 91 years (95% CI: 35.06, 36.58). The mean age of female inmates was 34.0 years (SD 9.0 years; range, 11.64-56.36 years). The number of sons was provided for 340 male inmates (mean, 2.47, [SD 1.96]; 95% CI: 2.26, 2.68). The mean number of sons of female inmates was 2.0 (SD 1.73; 95% CI: −2.30, 6.30). Time of incarceration was available for 706 inmates (95.3%) (mean, 31.33 months [SD 50.77 months]; 95% CI: 27.58, 35.08; range, 1-360 months). The mean treatment time was 56.79 months (SD 75.63 months; 95% CI: 51.86, 61.73 months) (Table 1).

### Association of HIV and co-infections

HIV-TB, OR 6.4 (95% CI: 3.5, 11.3); HIV-(HBV+HCV), OR 30.9 (95% CI: 19.1, 48.2); HIV-syphilis, OR 63.7 (95% CI: 41.4, 96.7). There was no association with others variables such as age, treatment, and time of incarceration. There was a statistical significance (*P*<0.001) for those with HIV acquiring co-infections.

### Prevalence and rates of inmates in prison unity

The Forest plot (Figure 2) shows the prevalence and coefficient intervals of infectious diseases in each unit analyzed. In 21 units, there was no relationship between the number of prisoners and the prevalence of infectious diseases. The best relationship was found in Presidente Venceslau, Florínea, Lucélia, and Presidente Bernardes. Figure 3 shows that the number of inmates by prison varied from low (66-500) in 4 prisons to high (1,501-2,500) in 12 prisons (42.9%). Four municipalities had 2 prisons and one had 3 prisons.

**Figure 3.**
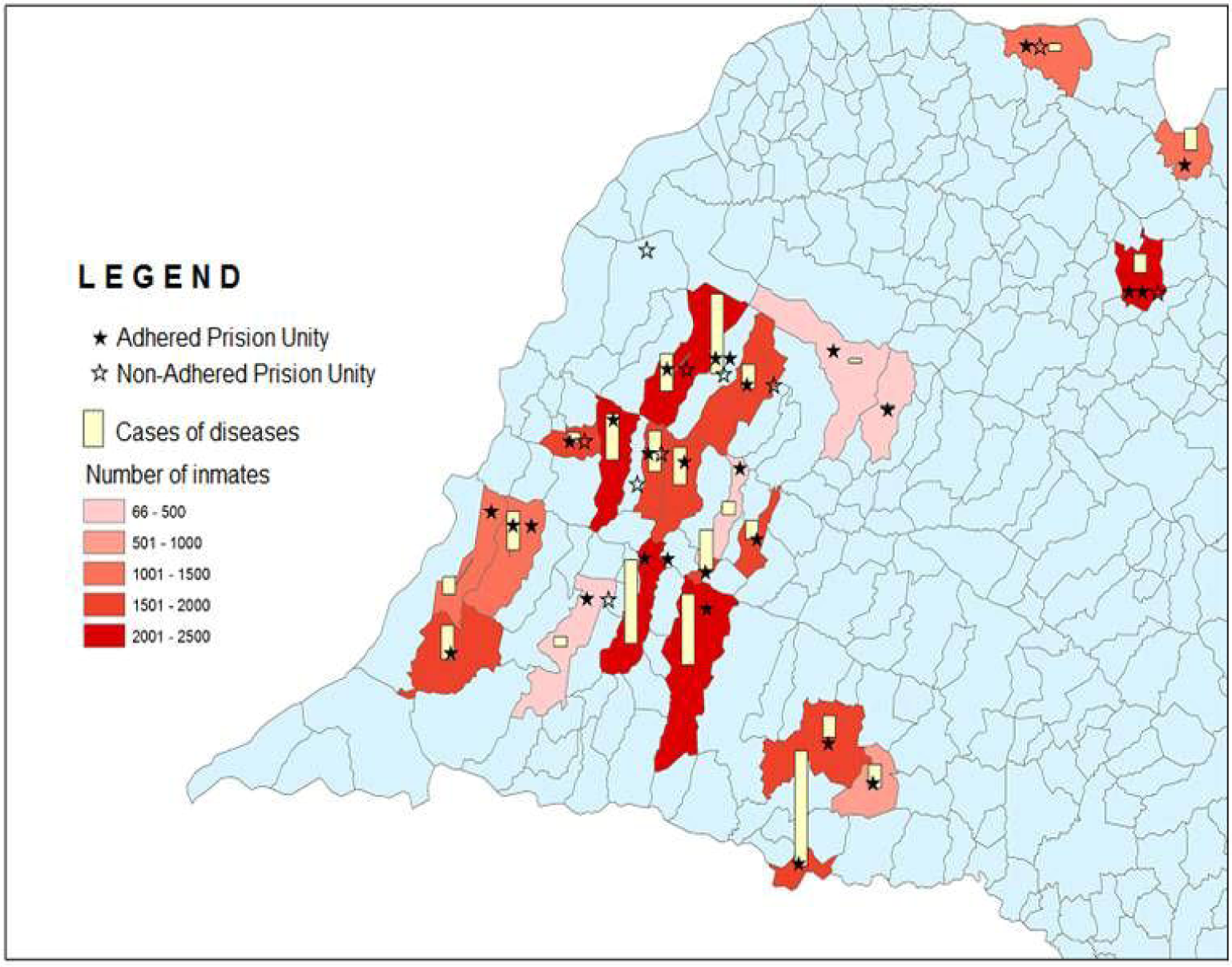
Geospatial location of prisons in municipalities in São Paulo state in the CROESTE domain. Bars represent the number of cases of infectious diseases in each prison. Colors represent the interval number of inmates in each municipality. Stars represent the number of prisons in each municipality. Current shape file databases and base maps were downloaded from the IBGE website.

### Geographic location of prisons and the highways network

Of 28 prisons, 18 (64,3%) were constructed in the western region near Presidente Prudente (number 18), the largest city with an airport, reference public hospital, Ambulatory of Medical Specialization service, and a public reference laboratory (Figure 1C). Strategically, most prisons were constructed beside radial highways: SP-300 Marechal Rondon is 346 miles long; SP-294 João Ribeiro de Barros is 295 miles long; SP-270 Raposo Tavares highway is 400 miles long. Transverse highways are important for the transit of inmates to Presidente Prudente and other large cities outside CROESTE. The most important is SP-563 Euclides de Oliveira Figueiredo highway (219 miles long).

### Population growth and Human Developing Index in municipalities with prisons

Table 2 shows the cities in which the prisons were built in the CROESTE domain (15). A higher than expected population growing was observed in 17 areas (47.2%). However, in 18 areas (50%), the population growth was less than expected and in one city, the population remained the same. Some cities experienced high growth, including Marabá Paulista (50.43%), Riolândia (65.94%), Lavínia (130.2%/137.8%), and Pracinha (178.0%); in Presidente Bernardes and Florínea, the population decreased in the period (−2.84% and − 11.63%, respectively). The presence of prisons did not change the HDI and no great variation in their values was found. A lower HDI was found in Marabá Paulista (0.68) located at Pontal of Paranapanema, one of the poorest regions in São Paulo state and a higher HDI was found in Presidente Prudente (0.81).

**Table 2.** Population Growth and Human Developing Index in Municipalities with Prisons in the Western and Northwestern Region of São Paulo State, Brazil

| Municipality | Date the Prison Opened | Estimated Population |  | Population Growth (%) | Human Development Index |
| --- | --- | --- | --- | --- | --- |
|  |  | In the Year the Prison Opened | 2018 |  |  |
| Andradina | 17 Dec 1998<sup>a</sup> | 54.440 | 57.112 | 4.91 | 0.779 |
| Araçatuba | 9 Apr 2001 | 171.289 | 195.874 | 14.35 | 0.788 |
| Assis | 7 Nov 1991 | 85.391 | 103.666 | 21.4 | 0.805 |
| Birigui | 26 Apr 2003 | 100.207 | 122.359 | 22.11 | 0.78 |
| Caiuá | 15 Apr 2005 | 4.725 | 5.800 | 22.75 | 0.697 |
| Dracena | 17 Dec 2001<sup>a</sup> | 40.576 | 46.536 | 14.69 | 0.776 |
| Flórida Paulista | 16 Mar 2005 | 10.226 | 14.486 | 41.66 | 0.715 |
| Florínea | 26 Feb 2016 | 2.778 | 2.699 | -2.84 | 0.713 |
| Irapuru | 15 Apr 2005<sup>a</sup> | 6.956 | 8.261 | 18.76 | 0.712 |
| Icém | 10 Feb 2017 | 8.112 | 8.181 | 0.85 | 0.72 |
| Junqueirópolis | 19 Oct 1998 | 15.609 | 20.524 | 31.49 | 0.745 |
| Lavínia | 15 Jan 2002<sup>a</sup> | 5.067 | 11.667 | 130.25 | 0.721 |
|  | 25 Jan 2006 | 4.905 |  | 137.86 |  |
|  | 25 Jan 2006 |  |  |  |  |
| Lucélia | 04 Dec 1998 | 16.888 | 21.604 | 27.93 | 0.752 |
| Marabá Paulista | 04 Feb 2005 | 3.827 | 5.757 | 50.43 | 0.677 |
| Martinópolis | 17 Mar 1999 | 22.484 | 26.289 | 16.92 | 0.721 |
| Mirandópolis | 08 Mar 1991 | 24.433 | 29.418 | 20.4 | 0.751 |
|  | 30 Apr 1993 | 24.898 |  | 18.15 |  |
| Oswaldo Cruz | 11 Mar 2002 | 29.802 | 32.754 | 9.91 | 0.762 |
| Pacaembu | 29 Sep 1998<sup>a</sup> | 11.138 | 14.130 | 26.86 | 0.725 |
|  | 12 Dec 2001 | 12.528 |  | 12.79 |  |
| Paraguaçu Paulista | 15 Jan 2002 | 40.834 | 45.455 | 11.37 | 0.762 |
| Pracinha | 23 Jan 2002 | 1.424 | 3.971 | 178.86 | 0.696 |
| Presidente Bernardes | 16 Nov 1990<sup>a</sup> <sup>b</sup> |  | 13.190 |  | 0.757 |
|  | 02 Apr 2002 | 14.926 |  | -11.63 |  |
| Presidente Prudente | 23 Apr 1990 <sup>b</sup> |  | 227.072 |  | 0.806 |
|  | 22 Feb 2002 | 194.173 |  | 16.94 |  |
| Presidente Venceslau | 05 Dec 1961 <sup>b</sup> |  | 39.448 | NA | 0.763 |
|  | 23 Oct 1999 | 35.327 |  | 11.67 |  |
| Riolândia | 15 Dec 1998<sup>a</sup> | 7.478 | 12.342 | 65.04 | 0.703 |
|  | 22 Nov 2013<sup>a</sup> | 11.429 |  | 7.99 |  |
| São José do Rio Preto | 24 Oct 2002<sup>a</sup> | 374.745 | 456.245 | 21.75 | 0.797 |
|  | 06 Aug 2004 | 398.079 |  | 14.61 |  |
|  | 28 Dec 2010 | 408.258 |  | 11.76 |  |
| Tupi Paulista | 16 Mar 2005 | 12.810 | 15.404 | 20.25 | 0.769 |
|  | 16 Aug 2011<sup>a</sup> | 14.344 |  | 7.39 |  |
| Valparaíso | 28 Sep 1998 | 18.371 | 26.130 | 42.24 | 0.725 |
|  | 17 Dec 2001<sup>a</sup> | 18.823 |  | 38.82 |  |

Abbreviation: NA, not applicable.
<sup>a</sup>Prison did not adhere to the study.
<sup>b</sup>Data not found.

## DISCUSSION

Most Brazilian studies on infectious diseases in prisons were conducted in just one or in a few units. [17-18] HIV was the most prevalent infectious disease diagnosed with prevalence rates of 0.68%, lower than 1.3% suggested by countrywide national data. [19] In the western region, all inmates diagnosed with HIV are treated and are followed by specialists in reference centers in Presidente Prudente. [20-21] Prevalence of 1.8% was detected in a prison in São Paulo state. [7] In a prospective study of 3,362 inmates from 12 prisons in the central-west region of Brazil in 2013, a prevalence of 1.6% was found. [22] Dolan et al. [23] reported that an estimated 10.2 million people are incarcerated worldwide and 3.8% are living with HIV. In Latin America, prevalence varies from 1.5% to 5%. [23]

TB ranked second with a prevalence of 0.7%, similar to 0.9% found in prisons countrywide. [23] Since the discovery of HIV, the concept of double-trouble for HIV-TB co-infection highlights the interlinked issues. Prisons act as incubators for these agents because they are associated with higher levels of infection than in the surrounding population. [23-24] In Brazil, The incidence coefficient of TB decreased from 42.7 cases per 100,000 inhabitants in 2001 to 34.2 in 2014. [11] Conversely, countrywide there was an overwhelming increase in TB rates in prisons. In a survey carried out for the national notification database, in 2009– 2014, TB cases among prisoners increased by 28.8%. [25] Corroborating these data, in the 12 prisons in Central-West, a prevalence of 0.7% was found at baseline and 1.8% 1 year later. [22] At the regional level, there are few data on São Paulo state. In 2008, an observational study on 2,435 inmates in one prison and one jail in the city of Guarulhos, in the metropolitan area of São Paulo, a prevalence of 0.83% was found. [26] Thus, the prevalence in these studies was similar to our data. In this study, HIV-positive patients were more likely to be co-infected with other diseases. Of 82 individuals who had a co-infection, only 1.08% did not present HIV. The OR for HIV patients co-infected with TB was 6.4. Worldwide, compared with the general population, people living with HIV/AIDS are 26 times more likely to develop active TB, and in Brazil, this risk is 28 times higher, quite different from the rates found in the studied population. [27] The data demonstrate the overwhelming need to increase the search for patients with HIV-TB co-infection in prisons in the CROESTE domain.

HBV and HCV are serious global public health problems characterized by chronic infections that may not show symptoms for a long period, sometimes years or decades. [28] In this study, the prevalence of HCV and HBV was 0.2% and 0.04%, respectively, lower than 0.6% found in prisons countrywide for both HBV and HCV. [12] The prevalence of HCV in inmates is similar to that in the general population (estimated 0.7% in 2017). [29] In 2013, 15.1% and 4.8% of the worldwide incarcerated population had HCV and chronic HBV infection. [23] In 2007, a cross-sectional study conducted in a prison in São Paulo reported that the prevalence of HBV and HCV was 21.0% and 5.3%, respectively, [7] similar to 18.9% and 6.1% obtained in women incarcerated in the largest prison in Goiás state, central Brazil. [30] Our rates are about 80-fold lower than the global and local prevalence of HBV and HCV. The association of people living with HIV co-infected with viral hepatitis showed an OR of 30.9. Countrywide, from 2007 to 2017, 9.4% of the total cases of HCV and 5.2% of cases of HBV were co-infected with HIV, different from 0.06% and 0.04% for HIV-HCV and HIV-HBV, respectively, found in this study. In Brazil, the risk of HIV-infected individuals acquiring HCV is 51 times higher than in the general population. [29]

Syphilis, a sexually transmitted infection (STI), is a genital ulcerative disease caused by *Treponema pallidum* and associated with significant complications if untreated. [31] In this study, syphilis ranked third with a prevalence of 0.2%, 2.5-fold lower than the national data for inmates (0.5%). [19] In São Paulo state, in 2003 the prevalence of *T. pallidum* in Ribeirão Preto Penitentiary was 3.0%. [32] In 2007, in a prospective study among 680 individuals in a prison in São Paulo city, the prevalence was 5.3%. [7] In a prospective study conducted in 12 prisons in Central-West Brazil, the prevalence was 9.4%. [22] In a retrospective analysis using a methodology similar to our study, at the Regional Prison of Santa Cruz do Sul, Rio Grande do Sul, a prevalence of 6% was found. [34] Our data are about 15- and 26-fold lower than in the studies conducted in São Paulo state, suggesting that similarly to HIV and viral hepatitis, the disease is underdiagnosed with serious implications for public health. Women visitors are at high risk for HIV/syphilis infection from their partners and most of them are not aware and underestimate their risk. [35] In this study, the OR for HIV patients co-infected with syphilis was 63.7, a higher OR among the co-infections. The countrywide prevalence of HIV-syphilis co-infection is not known, because there are no co-infection data in the notification and investigation records of acquired syphilis or HIV/AIDS in the Notification of Injury Information System (Sinan).

The lower prevalence rates of HIV, HBV, HCV, and syphilis found in this study compared with countrywide and São Paulo state may reflect omission and incorrect information given by the health care staff responsible for completing the questionnaire. Furthermore, others factors may be implicated, including: (i) deficient numbers of health care professionals and their continuous turnover, with discontinuation of the active search programs in the prisons; (ii) difficulties in assessing diagnosis and treatment because there are no doctors available in some units and inmates must go to hospitals, health care, or reference centers. In this scenario, a high security operation is necessary. This is also referred by health care workers from maximum security penitentiaries where it is necessary to follow the prisoner from their cell to the ward; (iii) inefficient communication from the prisons about diagnosis of infectious diseases. Information is lost when the inmate is transferred from one unit to another.

With respect to age group, 65.9% of the infected individuals were between 21 and 40 years old. It is well known that in this period, men and women are most susceptible to STIs. [17,36-37] In this study, 50.0% failed to complete elementary school, showing that low schooling is a risk factor for being arrested and acquiring infectious diseases. [17] Most inmates (76.9%) self-denominated as heterosexual; however, it is well known that for different reasons, there is promiscuity in prisons. This number is probably overestimated; many are bisexual, highlighted by the fact that countrywide, 60% of inmates had single civil status. [4] Men who have sex with men (MSM) are particularly vulnerable to STIs (11.9% in our study). [36]

The number who received treatment for the diseases investigated in this study was significantly higher than those who did not, mainly due to the Brazilian Ministry of Health’s TB and HIV Surveillance and Control Programs. In our study, 57.9% had been in prison previously, favoring the possibility of primary infection or reinfection, especially of TB. Worldwide, prison health care is currently deficient in the continuity of care. [23] In the region, a high number of prisons have great variation in the number of inmates, from just 66 to 2,500 individuals (Figure 3). The Forest plot shows that there was no relationship between the number of inmates and the number of infected individuals in 75% of the units (Figure 1). Although there is a protocol to be followed statewide for active search for infectious diseases in prisons, the diagnosis depends in part on the implementation and attitude of the health care team in each unity. The best relationships between the number of inmates and infected individuals were found in Presidente Venceslau, Florínea, Lucélia, and Presidente Bernardes prisons.

The main objectives of public authorities in constructing several prisons in the western and northwestern region were decentralization from São Paulo, to increase the regional economy and development, and to decrease migration of people from small- and medium-sized cities to large cities. In this aspect, higher than expected population growth was observed in 47.2% of the cities. If these interventions had not been implemented, this scenario would be even worse. We demonstrated that probably the region was chosen by public authorities due to availability of the highways network linking small, medium, and large cities and São Paulo. Most inmates were transferred from Carandiru and families living in São Paulo and in the metropolitan area must have easy access to prisons for visits. The increasing presence of these large buildings next to the highways has completely changed the landscape of the region.

Some shortcomings must be highlighted in our study. As a retrospective study, **s**ome important data cannot be measured and there was no selection of controls. There was a lack of commitment by many units in responding to the questionnaire, requiring several reminders. Long distances between prisons made access difficult and expensive.

## CONCLUSIONS

The prevalence of infectious diseases among inmates is lower than countrywide and the diagnosis depends in part on the implementation and attitude of the health care team in each unit, among a population characterized as young, heterosexual, and with previous imprisonment. Prisons have been constructed beside radial highways and can prevent migration of people from small- to large-sized cities. We suggest that improvements are necessary mainly in screening for infectious diseases in prisons in the western and northwestern region of São Paulo state. Results may be applied by public health authorities mainly in developing countries.

## Data Availability

This is laboratory and epidemiological data of prisoners who live mostly in maximum security prisons. Therefore, data cannot be made available for consultation

## ACKNOWLEDGMENTS

Roberto Medina and Denise Yukiko Tomokane for the support of data collection in each prison unit.

## AUTHOR AFFILIATIONS

Department of Post-Graduation, Master’s Degree in Health Sciences, Universidade do Oeste Paulista, Presidente Prudente, São Paulo, Brazil (Charlene Troiani do Nascimento, Danilo Zangirolami Pena, Rogério Giuffrida, Fernanda Nobre Bandeira Monteiro, Francisco Assis da Silva e Luiz Euribel Prestes Carneiro); Statistics Department, School of Sciences and Technology, São Paulo State University, Presidente Prudente Campus, Presidente Prudente, São Paulo, Brazil (Edilson Ferreira Flores).

## AUTHORS’ CONTRIBUTIONS

CTN, collection, tabulation, study design, and writing of the manuscript; DZP, tabulation; EFF, RG, FNBM, FAS, geospatial statistics; LEPC, drafting and critical review of the manuscript.

